# What do we know about smell and taste dysfunction by SARS-CoV-2. Predictive Value of the Venezuelan Olfactory test and RT-PCR analysis in viral infection diagnosis

**DOI:** 10.1101/2020.11.25.20238998

**Authors:** Rosalinda Pieruzzini, Carlos Ayala-Grosso, Jose de Jesus Navas, Wilneg Rodríguez, Nathalia Parra, Emily Luque, Aida Sánchez-Gago, Scarleth González, Alexandra Hagobian, Angeline Grullón, Karen Díaz, Mariano Morales, Melanie De Jesús, Sonia Peña, Luis Rodríguez, Luis L Peña, Ana Asaro, Magda Magris

## Abstract

**Background:** Smell and taste disorders are reported very frequently and at an early stage in the evolution of the infectious disease caused by the SARS-CoV-2. These symptoms could be sensitive and specific to establish the condition of the infection, and may suggest the flow of decisions as to further therapy. We asked whether smell and taste impairment are earlier and more sensitive symptoms than the RT-PCR molecular assays for SARS-CoV-2 detection.

**Methodology:** Subjects (N=275) with a probable COVID19 diagnosis were classified as follows: Symptomatic with chemosensory dysfunction, symptomatic without chemosensory dysfunction, and asymptomatic. Subjects received a general clinical and otorhinolaryngology examination. Evaluation of the chemosensory dysfunction was performed by means of the Venezuelan Olfactory Test and taste test. Nasal swabs and blood samples were analyzed by real-time polymerase chain reaction analysis (RT-PCR) and a rapid diagnostic test to detect the SARS-CoV-2 virus and antiviral antibodies, respectively. Patients had access to molecular tests and smell and taste evaluations every 3 to 5 days until they recovered.

**Results:** Out of 44 patients that were positive for RT-PCR SARS-CoV-2: 45.83% had smell and taste disorders and COVID19 symptoms, 23.61% did not have smell or taste disorders, but had COVID19 symptoms, and 30.55% were asymptomatic. Mild hyposmia and hypogeusia account for the most frequent chemosensory disorders accompanying common SARS-CoV-2 symptoms. Time to recover from the chemosensory dysfunction ranges from 3 to 14 days, up to a maximum of 5 weeks, while RT-PCR becomes negative after 21 days and up to 35 days in some cases. The Venezuelan Olfactory Test and taste test has a 61.68% positive predictive value, 45.83% sensitivity, and 68.7% specificity for SARS-CoV-2 infection.

**Conclusions:** Smell and taste disorders are frequent symptoms of SARS-CoV-2 infection, but not a significant predictor of the disease, as compared to the molecular RT-PCR test.

## INTRODUCTION

At the early stages of the SARS-CoV-2 infection, the smell and taste disorders seem to have some predictive value as to the evolution of the infection (1). Since the beginning of the pandemic, multiple studies have reported a very variable association between the smell and taste dysfunction and the presence of other symptoms that are characteristic of the coronavirus infection (1).

The first reports of a multicenter study that grouped clinical data from 12 hospitals, a population of 417 patients with mild to moderate COVID-19, list cough, myalgia and loss of appetite as the most common symptoms; while symptoms associated with otorhinolaryngology-related disorders were facial pain and nasal obstruction in at least 86% of the cases; and there were smell and taste disorders in 88% of the cases. The onset of the smell dysfunction was earlier than any other symptom in 11.8% of the cases (2).

The sudden loss of the sense of smell associated with an increasing number of cases of coronavirus was reported in the United Kingdom recently; however, the evidence seems to be circumstantial, because it was found as information provided in social media by subjects who had had this symptom in an isolated fashion. The authors reported 9 cases of sudden anosmia without other associated symptoms in the first 3 weeks of the onset of the coronavirus. It is worth mentioning that none of these patients was evaluated with a specific test to determine the presence of the disorder (3).

More recently, another study carried out at Hospital L. Sacco in Milan, Italy with 59 of the 88 patients of the hospital, established that 33.9% had at least one smell and/or taste disorder and 18.6% had both. In this group of patients, 20.3% had smell symptoms before they were admitted to the hospital and 13.5% during their stay (4). The presence of smell alterations associated with a viral infection is not new in otorhinolaryngology; many viruses may cause olfactory dysfunction due to an inflammatory process of the nasal mucosa and the development of rhinorrhea. Among the viral agents associated with these alterations are rhinovirus, parainfluenza, Epstein-Bar, and some coronavirus (4,5). However, the fact that olfactory dysfunction associated with SARS-CoV-2 is not essentially related to the onset of rhinorrhea and nasal obstruction suggests a different action mechanism. So far, the physiopathology of smell and taste disorders in the SARS-CoV-2 infection is still under scrutiny.

One of the most relevant evidences of the action mechanism of coronavirus was reported as a result of an investigation of the olfactory mucosa of mice and analysis of RNA sequences of humans. It found that the 2 genes that express the information for angiotensin-converting enzyme 2 (ACE2) and the host transmembrane serine protease family member II (TMPRSS2) receptors involved in the entry of COV-2 into the cell are expressed in the cells of the respiratory epithelium of the nasal cavity, support cells, Bowmanś glands, microvilli, and stem cells of the olfactory mucosa, but not in the olfactory sensory neurons. This suggests that the olfactory damage mechanism is non-neural in nature. As to the damage to the sense of taste, it appears to be directly on the taste receptor and as the result of the production of cytokines that irritate the trigeminal and glossopharyngeal nerves that transmit sensory signals to the central nervous system (6).

Netland et al. found that, in certain transgenic mice, there was an extensive replication of the virus in the brain, with certain regional differences, and that this brain infection, in turn, was an important factor in the aspiration pneumonia seen in some of the mice studied. A generalized neuronal infection appeared after intranasal inoculation, which suggests that the entryway for the infection could be through the olfactory bulb. This raises questions about the possible effects of coronavirus on patients infected with SARS-CoV-2 in the future (7).

A vast majority of initial reports on smell and taste dysfunction related to coronavirus infection were based on results from patient surveys, without corroborating the diagnosis through psychophysical evaluation of chemosensory dysfunction. Patients were not followed up systematically until recovery, nor was the relationship proven by means of diagnostic tests, such as the SARS-CoV-2 RT-PCR test, or rapid diagnostic tests. These factors may have led to an overestimation of the symptom as predictive factor of the disease. Similarly, using the smell test without adjusting it to the population under analysis could result in some false positives and, consequently, an overestimation of chemosensory dysfunction diagnostics.

All of the above motivated us to carry out an investigation the distinctive nature and main objective of which was to determine the predictive value of smell and taste tests in the diagnosis of SARS-CoV-2 infection, and its relationship with other diagnostic tests, such as SARS-CoV-2 RT-PCR test and the rapid diagnostic test, as well as the follow up and comprehensive observation of the patient’s recovery from the coronavirus infection. The objective of this study is to determine whether smell and taste disorders are an early and more sensitive biomarker than the RT-PCR test for diagnosing the SARS-CoV-2infection.

## MATERIALS AND METHODS

### Population

A sample of 275 individuals was selected from a population of 525 subjects examined between March and August 2020 (Table 1). The following were collected for all subjects at the beginning of the study protocol: 1. Full clinical record, including age, gender, epidemiologic background on the basis of travel and contact with positive cases, questions about suggestive respiratory signs and other symptoms, presence or not of smell and/or taste disorders. 2. General otorhinolaryngology (ORL) physical examination, Venezuelan Olfactory Test (VOT), and Basic Taste Test evaluation. 3. Nasopharyngeal swab for SARS-CoV-2 by RT-PCR molecular analysis and detection of SARS-CoV-2 antibodies by rapid diagnostic test (RDT).

**Table 1.** SARS-CoV-2 Group Definition

**+58 424 1367083**
| Table 1. SARS-CoV-2 Group Definition |  |  |
| --- | --- | --- |
| Symptomatic with smell and taste dysfunction by VOT (SyMVOTT+) | Symptomatic without smell and taste dysfunction by VOT (SyMVOTT-) | Asymptomatic (ASyM) |
| N=107 | N= 61 | N=107 |
| Subjects with general symptoms as described below: headache, cough, fever, general malaise, chest pain, dyspnea, myalgia, arthralgia, shivering, hyporexia, nausea, vomiting, diarrhea and any other symptom described in the literature that might be present and/or suggest a SARS-CoV-2 infection **and positive smell and taste results according to the VOT\* rating and positive smell and taste results in the VOT grading** | Subjects with general symptoms as described below: headache, cough, fever, general malaise, chest pain, dyspnea, myalgia, arthralgia, shivering, hyporexia, nausea, vomiting, diarrhea and any other symptom described in the literature that might be present and/or suggest a SARS-CoV-2 infection | Patients who did not present with any prior symptoms or signs associated with the coronavirus. |
| VOT: Venezuelan Olfactory Test |  |  |

In this study, patients were followed-up every 3 to 5 days, to verify their overall clinical condition. In addition, smell and taste tests were carried out, blood samples were drawn to perform RDTs, and nasopharyngeal swabs for RT-PCR tests, until they met the recovery criterion. The recovery criterion is met when one of the following conditions is fulfilled: 1.-The relative smell and taste test score is at the highest level and the RT-PCR test is negative. 2. - It has been established that there is no permanent smell or taste disorder.

### Clinical assessment

The clinical assessment of patients with probable COVID19 diagnostic followed the guideline suggested by the National Institute of Health of the United States of America (NIH-USA SARS-CoV-2 assessment guidelines). The level of organ compromise is defined as ***mild or moderate*** if individuals present with fever, cough, sore throat, general malaise, headache, myalgia, anosmia, ageusia, and diarrhea. When it is a ***mild*** condition, patients do not complain of shortness of breath, have more than 94% oxygen saturation, and there is no clinical evidence of respiratory disease, while in the ***Moderate*** there is clinical and imaging evidence of mild respiratory disease. The condition is defined as ***Severe*** in individuals with less than 94% oxygen saturation and respiratory rate at 30 respirations per minute in adults, in addition to more than 50% pulmonary infiltrates or a PaO2/FIO2 ratio lower than 300 mmHg. In the case of ***Critical*** disease, there is respiratory distress which requires mechanic ventilation, as well as multiple organ failure, septic shock and vasopressor treatment.

### Venezuelan Olfactory Test (VOT) and Taste Test

The VOT is a smell test that was adapted for Venezuela from the smell identification test of the University of Pennsylvania (UPSIT) (8). The Venezuelan short test is composed of 2 notebooks with 10 odorants that are commonly recognized by Venezuelans and which were validated during a prior exploratory study (9, 10). The substances are coffee, chocolate, baby cologne, scented talc, liquid detergent, coconut essence, powdered detergent, cinnamon, acetone, and rum. Every time an odorant is examined, the subject must identify the respective smell and discriminate it among the other substances offered and mentioned in addition to the odorant he is expected to identify.

On the basis of identification and discrimination of odors presented to patients, the VOT provides a relative grading scale as follows: Normosmia (8-10), mild hyposmia (6-7), moderate hyposmia (5-4), severe hyposmia (2-3), and anosmia (0-1).

The taste test consists in recognizing the 5 universally accepted basic tastes, to wit: sweet, salty, sour, bitter, and umami (11). One (1) cc of a sample of each taste is placed on the anterior third of the tongue of the patient who has his eyes closed, and after 10 seconds of period, an identification of the sample may occur. If the patient cannot recognize the tastes, he is diagnosed with ageusia, and hypogeusia if the patient recognized up to 4 tastes.

### Rapid Diagnostic Test (RDT) to Determine Antibodies Generated to SARS-COV-2 virus antigens

Diagnostic tests N° IFU-COVID3-01 (Nhui deep blue Medical Technology co. Batch: 20200307) were used to determine IgG/IgM antibodies. This kit uses a recombinant SARS-CoV-2 antigen conjugated with colloidal gold, which can interact with antibodies circulating in blood or in a serum or plasma preparation. The sample is placed on a nitrocellulose (NC) membrane that has been prepared with rat anti-IgM antibodies and human IgG antibodies, and sheep anti-rat polyclonal antibodies. When the sample diffuses in the membrane it may interact with the IgM antibodies adsorbed in it and an aggregate is produced with the coronavirus antigen marked with colloidal gold. This, in turn, is captured by the rat anti-IgM and produces a colored line. If the sample contains IgG antibodies, it forms a compound with the antigen marked with colloidal gold, this compound is captured by the rat anti-human IgG antibody and produces a colored line. When samples contain IgM and IgG at the same time, there will be 2 lines. When there are no IgM or IgG antibodies in the sample, there is only the quality control line and the result is negative.

### Sample Collection for RT-PCR Testing for SARS-CoV-2 detection

Samples were obtained from patients by nasopharyngeal swab strictly following the biosecurity protocol. Then, they were placed in YOCON viral transport medium, batch Y25200101, and kept at a temperature between 2 and 8° C until they reached the reference laboratory. Samples collected for RT-PCR testing for SARS-CoV-2 detection were processed by the Virology Service of the “Rafael Rangel” National Hygiene Institute of the Ministry of the People’s Power for Health in Caracas, Venezuela.

### Ethical Approval

Collection and analysis of data were approved by the Bioethical committee of the “Carlos Arvelo” University Military Hospital of Caracas. All studies were conducted in compliance with the Declaration of Helsinki, and all participants signed an informed consent. The current study included participants for which there was full information on multiple SARS-CoV-2 measures and key outcomes, including psychosocial factors, chronic medical conditions, and socio-demographic factors.

### Statistical Analysis

Descriptive statistics was used in the analysis by age group and type of chemosensory impairment. The predictive value, sensitivity and specificity of diagnostic tests used were calculated on the basis of the Wilson score method using the OpenEpi, version 3 software. Diagnostic Test Open code Calculator.

*Χ*^2^-Square tests were used to find the most frequent type of symptom in the SARS-CoV-2 infection. We asked whether there is a relationship between the SARS-CoV-2 infection and the presence or absence of symptoms; and to evaluate the relationship among type of olfactory disorder, age and gender. In the case of the association between two variables, when the result of the *Χ*^2^-Square test was positive, that is to say, the variables were dependent on, or associated to one another, a standardized residuals analysis was carried out. This analysis allows to determine in a significant manner which cell or frequency contributed more to the rejection of the null hypothesis in the *Χ*^2^-Square test. Additionally, it allows to find out which cells deviated significantly from the expected value. Any deviation value higher than ±1.96 from the normal distribution is considered significant.

Pearson’s *Χ*^2^-Square test and Fisher’s exact test were used to evaluate between-group differences in two categorical variables.

*Χ*^2^-Square tests and standardized residuals analyses were carried out in R with the chisq.test function of the stats. package (12).

## RESULTS

The most frequent age group among study subjects was the 25 - 38 years old, and there were more male (54.90 %) than female (45.09 %) subjects; average age of the population was 33.63 ± 5 years old (Table 2). In Venezuela, the first possible SARS-CoV-2 infection cases were reported in March (Figure 1). Initially, the disease was observed in a group of 28 patients with smell and taste disorders, although only 1 of them was confirmed positive for SARS-CoV-2 by means of the RT-PCR test. Then, starting in May, the number of individuals positive for SARS-CoV-2 was increasing in parallel with the onset of smell and taste disorders; and finally, in the month of August, a significant number of patients with smell and taste disorders also tested positive for coronavirus infection by means of the RT-PCR test.

**Table 2.** Distribution by Age and Gender

| Table 2. Distribution by Age and Gender |  |  |  |  |
| --- | --- | --- | --- | --- |
|  | Males |  | Females |  |
|  | 151 |  | 124 |  |
| Age Range | N | % | N | % |
| 18-24 | 27 | 9.81 | 16 | 5.81 |
| 25-31 | 41 | 14.90 | 32 | 11.63 |
| 32-38 | 31 | 11.27 | 28 | 10.18 |
| 39-45 | 15 | 5.45 | 19 | 6.90 |
| 46-52 | 16 | 5.81 | 9 | 3.27 |
| 53-59 | 17 | 6.18 | 10 | 3.63 |
| 60-66 | 4 | 1.45 | 10 | 3.63 |

**Figure 1.**
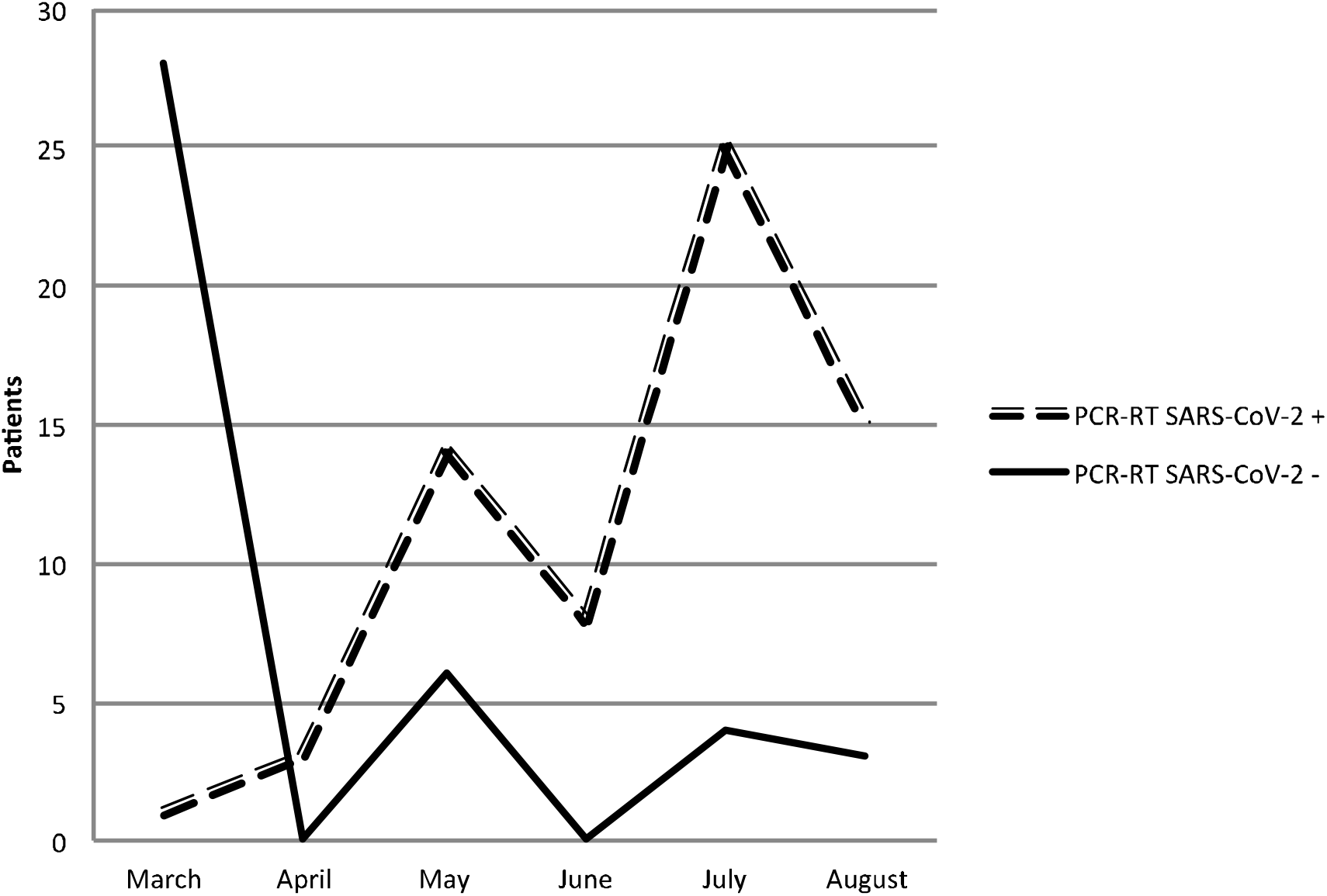
Time course of number of cases RT-PCR+ and RT-PCR-along months of COVID19. The broken line represents the patients with smell and taste disorders who were positive for COVID-19 by PCR-RT. An increase is observed since May with a maximum in July-August. The solid line represents patients with chemosensory disorders without being positive for COVID-19.

The observation of a higher incidence of olfactory and taste disorders together with a positive molecular diagnostic suggested that the dysfunction of these senses could happen before or to a greater extent than the expression of viral genes evidenced by the RT-PCR test, and this could imply that the physiology of the chemosensory system is more susceptible to SARS-CoV-2 infection.

To assess this observation, molecular tests were performed on the 275 subjects who had gotten smell and taste tests. It was found that 144 subjects were RT-PCR+ for SARS-CoV-2 while 131 were RT-PCR- for SARS-CoV-2. In the RT-PCR+ group, there were more SyMVOTT+ (66/144; 45.83%) subjects than SyMVOTT-(34/144; 23.61%) or ASYM (44/144; 30.55%). In contrast, among the 131 patients that tested negative for SARS-CoV-2 in the RT-PCR test, there were more SyMVOTT+ (41/131; 31.30%) subjects than SyMVOTT-(27/131; 20.61%), but less than the ASYM (63/131; 48.10%) (Table 3). These results suggest that there are more individuals with olfactory and taste disorders among the COVID19 symptomatic population.

**Table 3.** PCR and PDR Testing

| Table 3. PCR and PDR Testing |  |  |  |  |  |  |  |  |
| --- | --- | --- | --- | --- | --- | --- | --- | --- |
|  | PCR+<br>/PDR+ | PCR+<br>/PDR- | PCR+ |  | PCR-<br>/PDR- | PCR-<br>/PDR+ | PCR- |  |
|  | N=144 |  |  | % | N=131 |  |  | % |
| SyVOTT+ | 16 | 50 | **66** | **45.83** | 38 | 3 | **41** | **31.30** |
| SyVOTT- | 14 | 20 | **34** | **23.61** | 24 | 3 | **27** | **20.61** |
| ASyM | 17 | 27 | **44** | **30.55** | 38 | 25 | **63** | **48.10** |
| $\chi^2=9.19$, gl = 2, p < 0.05 | | | | | | | | |

The higher number of RT-PCR+ patients with and without olfactory and taste disorders, and with symptoms of the infection indicates that the relative effectiveness of RT-PCR tests in confirming the COVID19 infection is 75%. In contrast, if the large number of PDR-(197/275; 71.63%) individuals in the whole sample is taken into account, the relative effectiveness of this test is 28%. (Table 3). This confirms the little reliability of PDR tests for diagnosing coronavirus infection.

The frequency of subjects in the SyMVOTT+, SyMVOTT- and ASYM groups as a function of the presence of chemosensory symptoms and the positive molecular test results was heterogeneous (X^2^ = 9.19, gl = 2, p < 0.05), which suggests that the type of symptoms and their association with chemosensory dysfunction is different in the case of SARS-CoV-2 infection. This is consistent with what was observed in the analysis of standardized residuals, since, in the SARS-CoV-2 infection, the chemosensory dysfunction is very frequent and, in this sample, very few subjects that presented with the disease were asymptomatic (Table 4.1, Figure 1). On the other hand, olfactory and taste disorders are infrequent in the general population (Tables 3 and 3.1).

**Tabla 3.1.**
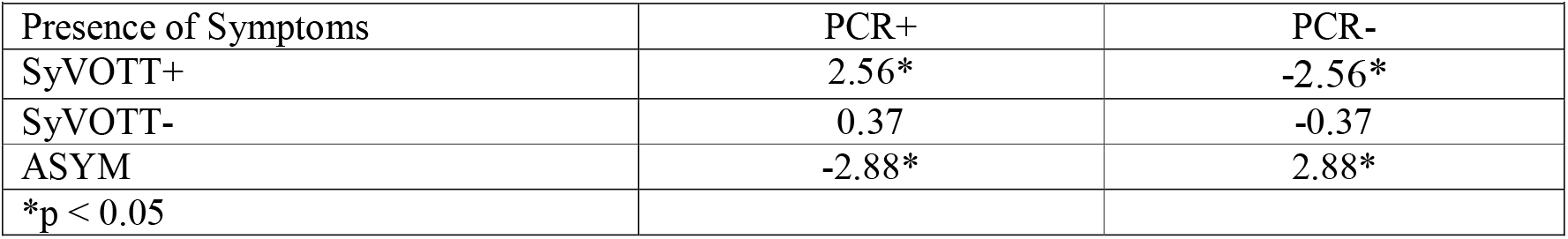
Standardized Residuals for Presence or Absence of Symptoms and SARS-CoV-2 Infection

|  |  |  |
| --- | --- | --- |
| Presence of Symptoms | PCR+ | PCR- |
| SyVOTT+ | 2.56* | -2.56* |
| SyVOTT- | 0.37 | -0.37 |
| ASYM | -2.88* | 2.88* |
| *p < 0.05 |  |  |

**Table 4.** Predictive Value of VOT and PCR for Diagnosing SARS-CoV-2 Infection

|  |  |  |
| --- | --- | --- |
|  | RT-PCR+ | RT-PCR- |
|  | N= 144 | N= 131 |
| SyVOTT+ | **66** | **41** |
| ASyM + SyVOTT- | **78** | **90** |
| $\chi^2$ 13.796, gl = 2, P < 0.05 | | |

### Predictive Value of VOTT

When the frequency of patients with olfactory and taste disorders that were diagnosed with the VOTT was compared to the RT-PCR tests positive results, the positive predictive value of the VOTT was 61.68%, while the negative predictive value was 53.57%. These results suggested that VOTT+ patients had a 0.62 probability of being RT-PCR+ for SARS-CoV-2. On the other hand, the sensitivity and specificity for COVID-19 diagnosis of the Venezuelan Olfactory Test and basic taste test was 45.83% and 68.7%, respectively (Table 4). These findings supported that these tests allow to detect olfactory and taste disorders in 46% of healthy individuals and in 69% of patients who were also clinically SARS-CoV-2 positive.

### SARS-CoV-2 Affectation and Chemosensory dysfunction

Coronavirus infection produces different degrees of affectation, as described in the methods. In this research, it was observed that 98.48 % of the 66 subjects (65/66) with chemosensory impairment (VOTT+) had mild clinical signs and 1.51% (1/66) moderate signs (Table 5).

**Table 5.**
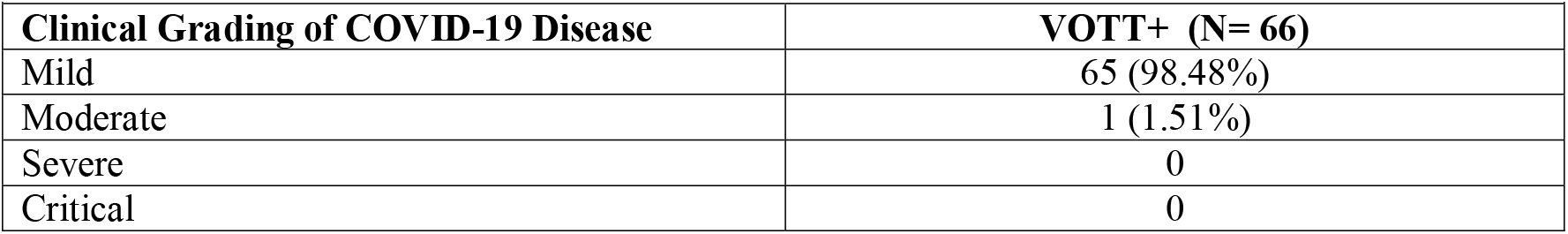
Association between Clinical Grading of SARS-CoV-2 Disease and a Positive Venezuelan Olfactory Test and Taste Test (VOTT+)

|  |  |
| --- | --- |
| Clinical Grading of COVID-19 Disease | VOTT+ (N= 66) |
| Mild | 65 (98.48%) |
| Moderate | 1 (1.51%) |
| Severe | 0 |
| Critical | 0 |

### Characteristics of VOTT+, SyVOTT +, and SyVOTT-Patients

Clinical signs as a consequence of SARS-CoV-2 infection are quite variable. The number of SARS-CoV-2 infected patients presenting with chemosensory disorder only (VOTT+) was 18/97 (18.55%). In contrast, there were 48/97 (49.5%) SyVOTT+ according to standardized residuals analysis P < 0.05; (Table 6).

**Table 6.**
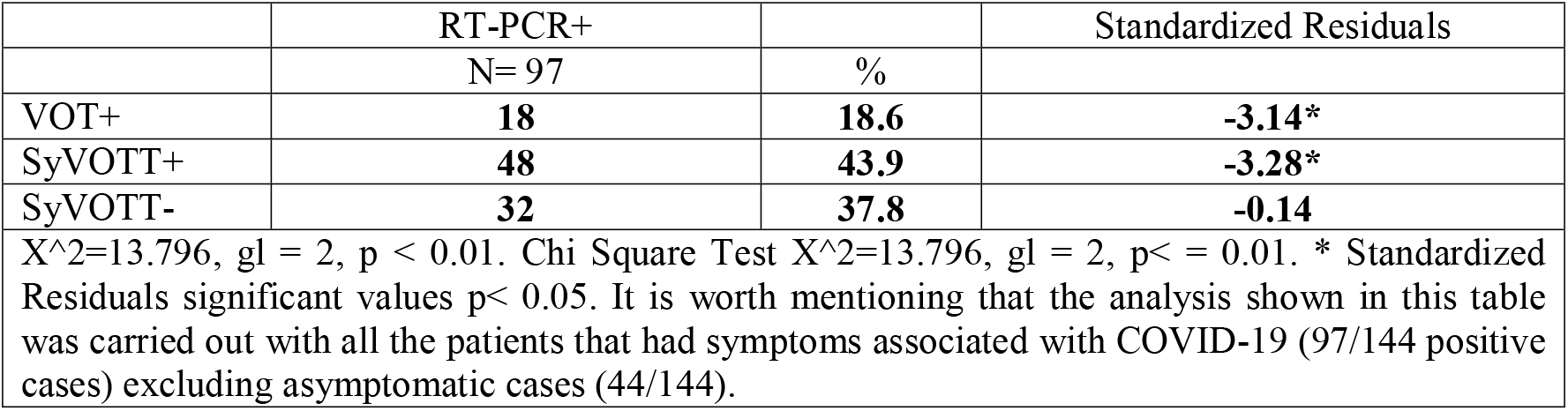
General Symptoms and Chemosensory Dysfunction in SARS-CoV-2 Infection

The group of VOTT+ patients without any other accompanying symptom (no concomitants), who referred olfactory and taste disorder as their only symptom before the molecular diagnostic and before being admitted to the hospital was 18/97 (18.55%) (Table 7). When differentiated on the basis of the type of chemosensory dysfunction, 7 presented with olfactory disorder only, 4 of them anosmia, and 3 with mild hyposmia. Combined smell and taste disorders at various degrees were present in 10 subjects, and only 1 had hypogeusia. All these findings suggest a disorder variability that may indicate the presence of coronavirus infection and not only the anosmia or the ageusia. In this group of patients, chemosensory disorders were an early biomarker of the coronavirus disease.

**Table 7.** VOT+ and taste + by Age and Gender

|  | Males |  |  | Females |  |  |
| --- | --- | --- | --- | --- | --- | --- |
|  | Age Range (years old) |  |  | Age Range (years old) |  |  |
|  | 18-35 | 36-51 | >52 | 18-35 | 36-51 | >52 |
| VOTT* | % |  |  | % |  |  |
| Anosmic | 0 | 3.0 | 1.5 | 1.5 | 3.0 | 3.0 |
| Mild Hyposmia | 7.6 | 1.5 | 1.5 | 7.6 | 6.1 | 1.5 |
| Moderate Hyposmia | 3.0 | 0 | 0 | 1.5 | 0 | 1.5 |
| Severe Hyposmia | 1.5 | 0 | 0 | 1.5 | 3.0 | 0 |
| Hypogeusia | 6.5 | 0 | 1.5 | 1.5 | 0 | 0 |
| Both | 7.6 | 6.0 | 4.5 | 13.6 | 3.0 | 6.1 |

VOTT\* : Venezuelan Olfactory Test and Taste Grading

Chemosensory dysfunction was a symptom in 31.25% of the subjects of the SyVOTT+ group before being admitted to the hospital. In contrast, 68.75% of the patients of the sample that did not declare the disorder before being admitted to the hospital, were positive for olfactory or taste disorder when the VOT and taste test were used. This confirms the need to use a standardized objective taste for detecting the chemosensory disorder during the hospital stay. On the other hand, the onset of the chemosensory dysfunction in the SyVOTT+ group occurred between the 3^rd^ and 5^th^ day in 70% of the cases.

Other associated symptoms in the SyVOTT+ group vary. Among them, headache, myalgia, arthralgia, shivering, odynophagia, and hyporexia in 48% of patients; fever, headache, and general malaise in 31%, dry cough and chest pain in 8% dyspnea, and only fever or myalgia in 8% and 2%, respectively.

Similarly, in the SyVOTT-patients, which accounted for 32.98% of the sample, there was fever, headache, myalgia, arthralgia, and shivering in 46.87% of them; headache, arthralgia and myalgia in 15%, as well as chest pain, dyspnea in 15%, rhinorrhea, fever, nasal congestion, and dysphonia in 12.5%; and cough, general malaise, fever, and shivering in 9%. As relates the variety of symptoms in SyVOTT+ and SyVOTT-groups, no differences were observed in the frequency of presentation that would warrant an additional classification in the SARS-CoV-2 infection.

### Age, Gender and VOTT

In this study, when the presence of chemosensory dysfunction in the subjects of the sample was taken into account, the frequency was similar among them regardless gender or age (Table 7). Furthermore, smell and taste disorders combined were present in 40.90% of VOTT+ patients. The severity of the disorder is anosmia and ageusia in 30% of the cases, while mild hyposmia and hypogeusia were observed in 59.25% of the cases. The olfactory disorder alone, was present as anosmia (12.12%), severe hyposmia 6.06%, mild hyposmia 25.75%, and moderate hyposmia (6.06%). It may be concluded from the analysis of the sample that the presence of both disorders is the prevailing clinical sign, and of those two, the most frequent olfactory disorder is mild hyposmia (32 patients). At the same time, anosmia was the second olfactory disorder in terms of frequency (17 patients). In contrast, hypogeusia as the only clinical sign is not frequent in coronavirus infection.

### Time Course of SARS-CoV-2 Grading by Means of VOT

The assessment of chemosensory dysfunction as a function of time by means of the Venezuela olfactory test allowed to establish a group of subjects with anosmia 11/31 (35.48%) and another with mild hyposmia 15/31 (48.4%) on day 1. The number of individuals positive for mild hyposmia and normosmia in the VOT increased in frequency with time, as follows: day 5, 8/10 (80%); day 7, 17/19 (89.5%); day 10, 8/8 (100%); day 14 8/10 (80%); and day 20, 15/16 (93.75%). Between day 5 and day 20, the sense of smell of 66/142 (46.47%) subjects had been restored (normosmia), as evidenced by the VOT test. The average recovery interval was 8 to 10 days, with a minimum of 3 days and a maximum of 20 (Figures 2A and 2B). Only one patient did not recover the sense of smell during the time of the study.

**Figure 2.**
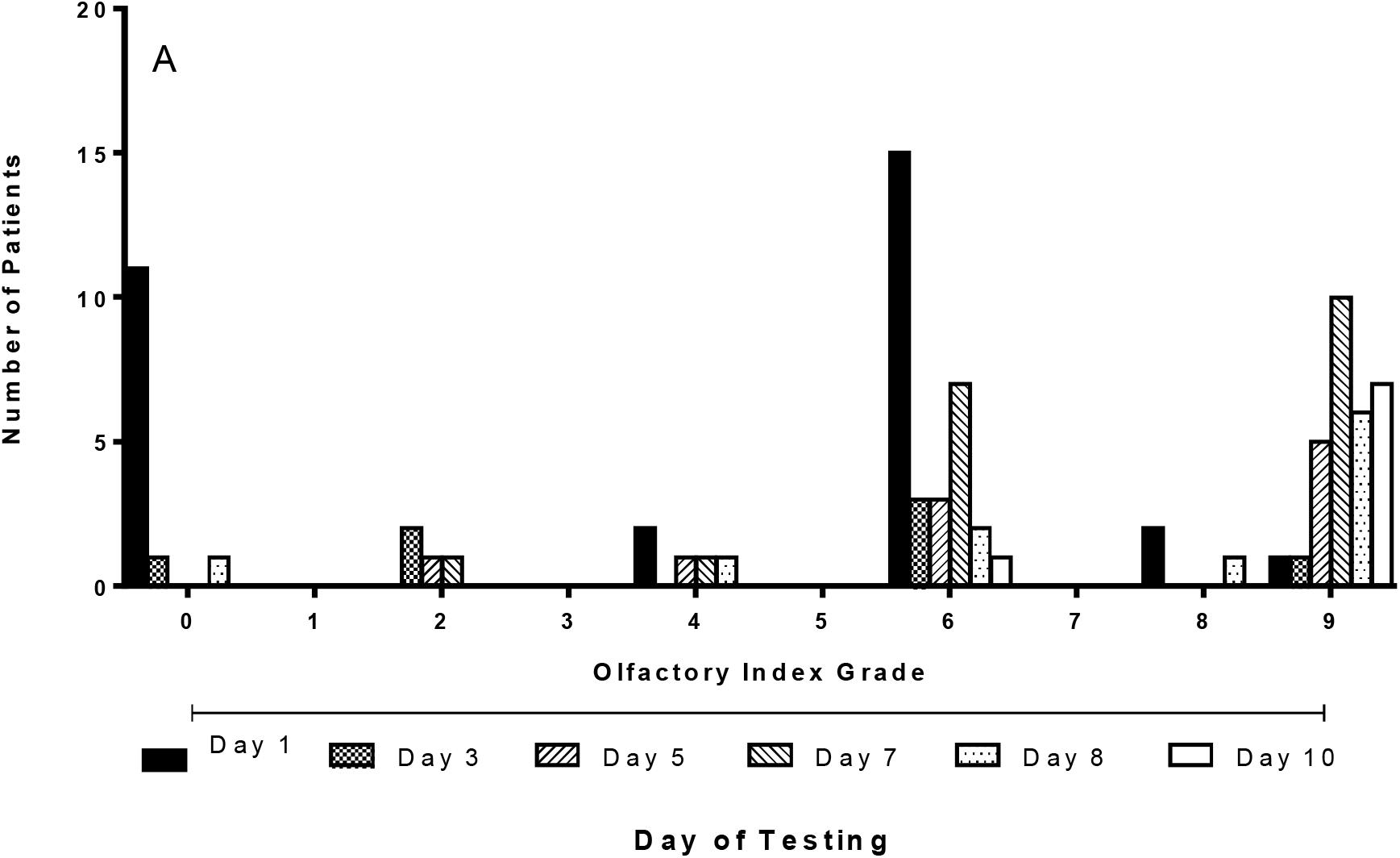
Olfactory impairment and evolution of SARS-CoV-2 infection. Venezuelan Olfactory Test was performed until Olfactory Index Grading reached normosmic value (8-10). A. Each bar represents number of subjects at specific grade of olfaction. Bar symbol represents day of testing (Day 1 -10). B. Each bar represents number of subjects at specific grade of olfaction. Bar symbol represents day of testing (Day 12 - 20)

### Time Course of SARS-CoV-2 Grading by Means of RT-PCR Test

The frequency analysis of the RT-PCR molecular test for SARS-CoV-2 was carried out for all individuals of the protocol in a systematic manner from day 1 to day 34 post infection. The viral gene expression was very frequent from day 1 to day 7, as it was present in 47/78 (60.25%) subjects; it decreased between day 10 and 14, and from day 16 to day 20, when it was evident in 15/78 (19.23%) and 11/78 (14.10%) subjects, respectively. Only 3 individuals had a positive RT-PCR test result on day 26 and only one did so until day 30. The frequency of VOTT+RT-PCR+ 66/275, and VOTT-RT-PCR+ 78/275 individuals is different from that of VOTT+RT-PCR-41/275, VOTT-RT-PCR-90/275 subjects (Fisherś exact test P < 0.0184). The frequency of RT-PCR+ molecular tests is higher between days 1 and 14 and lower between day 16 and 24. It was observed that the maximum time interval for viral gene expression was 21 days (Figure 3).

**Figure 3.**
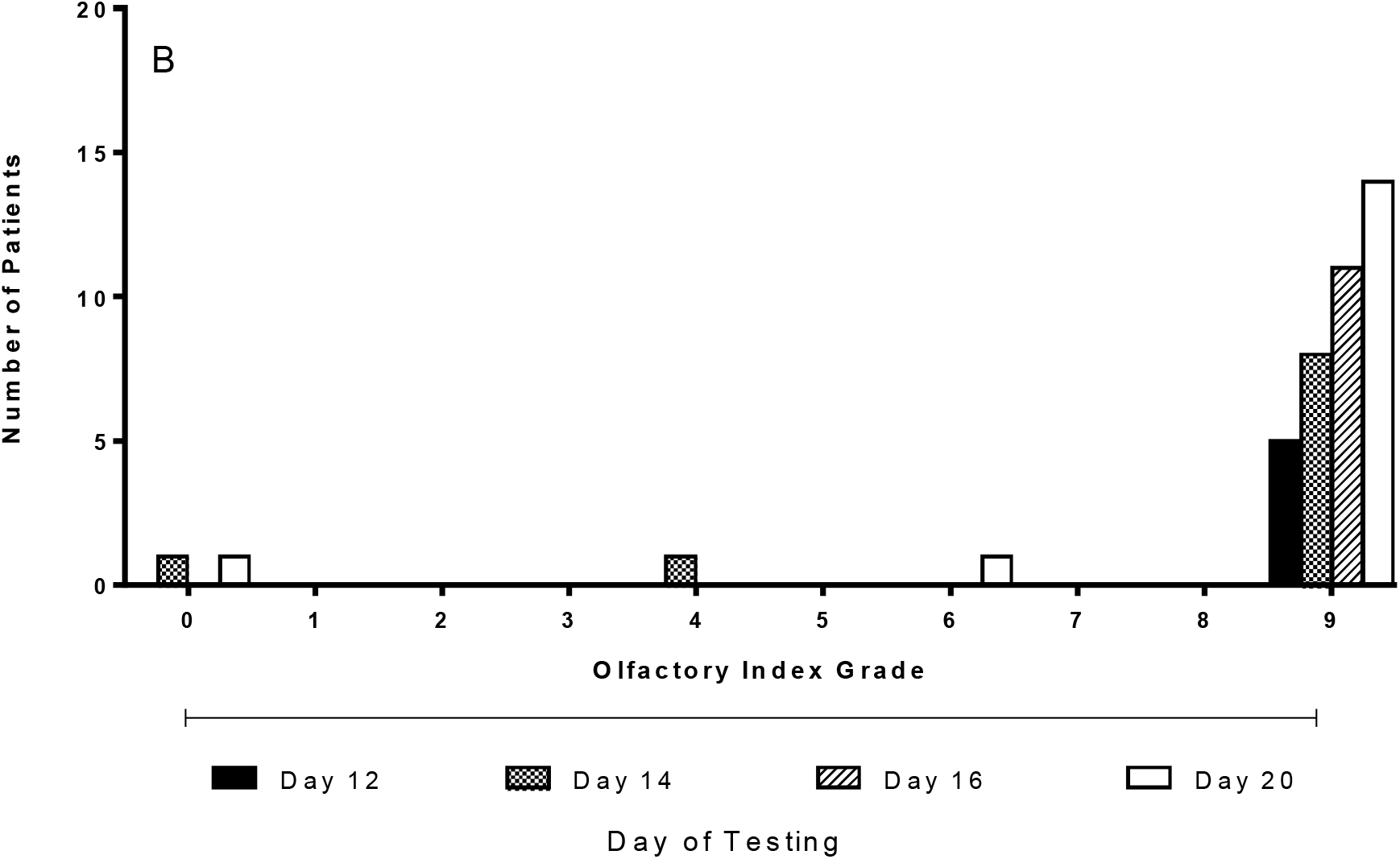
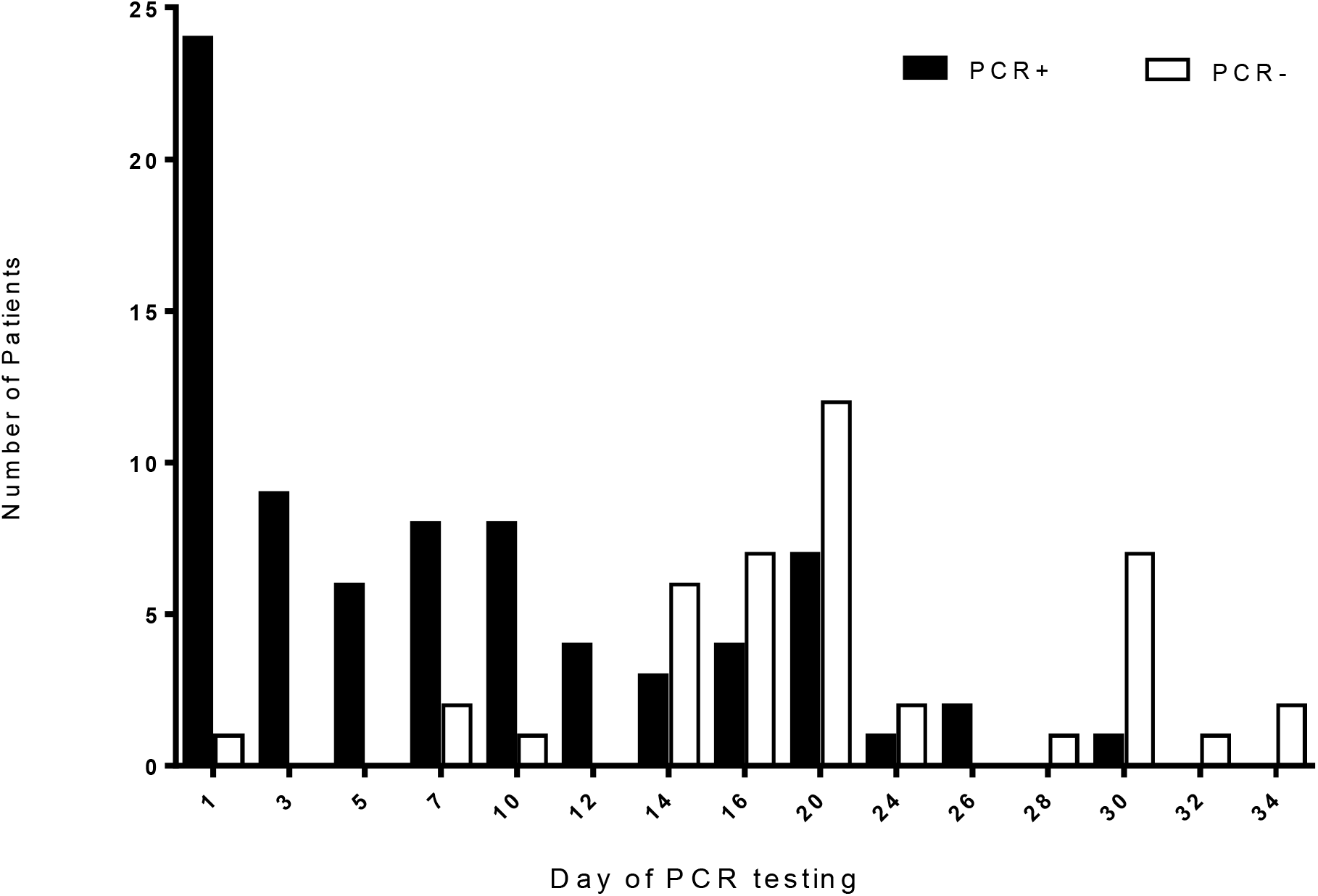
Evolution of RT-PCR molecular analisis under SARS-CoV-2 infection. RT-PCR molecular analysis was performed multiple times to each patient until testing become negative. Each bar represent the number of patients RT-PCR+ (black) and RT-PCR- (white) through time (days of PCR testing).

## DISCUSSION

The recent onset of this SARS-CoV-2 zoonosis in human populations has led to a significant number of studies aimed at understanding the variety of symptoms as part of the etiology of the disease. In the Venezuelan population, the time course of COVID19 has been lagging behind that of other countries, and this may have given time to establish treatment guidelines as a function of the expression of the disease. This study have evaluated the predictive value of a standardized smell test for the unbiased assessment of the sense of smell and taste in individuals with respect to a molecular diagnostic of SARS-CoV-2 infection by RT-PCR test. This is an original and innovative study, as it establishes in an objective and recurrent evaluation, by means of a smell test that is standardized for Venezuela and a classical taste test, the evolution of the chemosensory disorder of subjects until the sense of smell and taste are fully restored. Additionally, the viral genes analysis by means of the SARS-CoV-2 RT-PCR test and of the anti-coronavirus antibodies was carried out in symptomatic and asymptomatic patients until these had fully recovered.

Clinical signs vary in patients with coronavirus infection, and the severity of the disease is determined by shortness of breath, oxygen saturation, pulmonary infiltrates, multiple organ failure, septic shock, and the need to use vasopressors, as defined by the National Institute of Health of the United States of America (NIH-USA SARS-CoV-2 assessment guidelines).

Anosmia and ageusia were otorhinolaryngological and neurological signs of the coronavirus disease. Currently, the physiopathology of anosmia due to COVID-19 is apparently better understood; however, there is still the question why patients with moderate to severe COVID-19 infection have less olfactory affectation. Findings of this study show that 98% of patients with smell and taste disorders had mild COVID19. These results are in agreement with a recent report of self-described olfactory dysfunction in mild forms of SARS-CoV-2 infection that did not require hospitalization (13). For instance, the chemosensory dysfunction established in this study by means of self-evaluation underestimates the incidence of this symptom; therefore it would be advisable to examine whether anosmia is less prevalent in more severe forms of the infection and, consequently, the mechanism that leads to the physiopathological process that gives rise to this affectation.

The definition of a mild or severe form of SARS-CoV-2 infection may imply there is an immune response with more or less contention power thus leading to a milder or more serious viral infection, respectively. This response in the entryway of the virus may also imply that the progression of the infection depends on the time taken by the virus to move towards the upper or lower airways. Therefore, it is possible that, in the mild forms of the coronavirus infection, a more intense and faster immune response will produce more local inflammation that evolves into inflammatory processes involving the neuroepithelium and olfactory bulb. In contrast, in low intensity late immune responses, patients present with mild symptoms in ears, nose and throat; but later, these could also involve higher and lower airways with respiratory compromise (14, 15).

These two hypotheses have not been experimentally corroborated, because results show that patients with severe or critical forms have a higher immunoglobulin concentration in the serum and nasal secretions than patients with mild forms; which additionally makes sense, because the severe respiratory compromise has been justified as being a cytokine storm mechanism.

Since the beginning of the pandemic, COVID-19 symptoms reports have been about the respiratory tract affectation, with serious and fatal complications in a certain percentage of cases. The first reports involving a nervous system affectation were associated to a sudden decrease in the sense of smell, sometimes accompanied by a decrease in the sense of taste, which patients recovered from regardless whether they had a positive infection outcome or not. Initial reports were based on telephone interviews or online self-assessments by means of a questionnaire. As a result, the incidence of the chemosensory disorder was 19.4% (2); 64% (16); and 85.6% (15).

These findings confirmed the initial observation of Lechien et al. who were the first to mention the sudden loss of the sense of smell in SARS-CoV-2 infected patients. Subsequent reports expanded on the vision of this author and confirmed the association between the infection by this virus and the chemosensory disorder. Self-assessment of the senses of smell and taste has been proposed as an index of SARS-CoV-2 transmission that could be of value for projecting the need of hospital care units required for compromised patients (17). However, as it is commonly the case in these smell and taste self-assessment studies, sensitivity and specificity are compromised.

Applying standardized tests to SARS-CoV-2 infected patients has allowed to determine the level of involvement of the sensory function in the progression of the viral pathology. In a previous study, an UPSIT test for the Persian population applied to a reduced number of patients in Iran established that the olfactory disorder is variable, as there was a reduced rate of anosmic subjects (25%), while the largest percentage had moderate and severe microsmia (60%). Therefore, it was suggested that this degree of sensitivity was not enough to be able to consider the smell test as an indicator of the progression of the infection (18).

The senses of smell and taste form a physiological system that is affected by COVID-19; however, the degree of affectation seems to be mild. This has been shown by the studies carried out using as objective evaluator the chemosensory test of the Clinical Research Center of Connecticut, which has shown, just like Moein et al., findings, that a very small number were anosmic, while most had moderate and mild hyposmia (80%). At the same time, the taste test established that the taste affectation was mild to moderate. All patients maintained a normal degree of discrimination and a subjective overall recovery of 66%; but up to 88% presented with a certain degree of chemosensory disorder (19).

In contrast with these results, in this study, the results of several smell and taste tests showed the progressiveness of the disorder and time to recover from the onset of the infection. Contrary to what was seen in prior studies, in our sample, mild hyposmia and anosmia accounted for the highest share with a progressive recovery. The onset of the infection and its evolution allowed to make an evaluation until the recovery of the patient.

Evidence of SARS-CoV-2 infection and its demonstration by virtue of new technologies have given rise to molecular tests with variable sensitivity and specificity. Findings suggesting an asymptomatic infection in significantly frequent cases of negative RT-PCR test results imply several possibilities, which include that individuals have either not been exposed to the infection or they received a reduced viral load. At the same time, the characteristics of the sample, as a result of collection, storage, preservation, and processing, could also contribute to the high number of negative results (20,21).

The time course of the infection, and the effectiveness of molecular tests to prove the infection, suggest an order in the events of infection progression that range from 5 to 7 days for the first clinical signs to become evident. The sensitivity of the molecular diagnosis increases in a delayed manner, as a function of the increase in the concentration of viral particles that could reach a maximum level on the 7^th^ day and up to the following 14 days, to then decrease gradually as a result of the immune response of the patient.

In contrast, in this study, the RT-PCR molecular test leads to suggest a minimum time of 1 day from the date of admission of the patient to the protocol and up to 14 days. This is the first study that established a multiple system for following up the progression of the chemosensory disorder and detecting the infection; be it by means of the molecular test or the SARS-CoV-2-specific antibodies detection test. The Venezuelan olfactory test is a tool that has been validated taking into consideration the characteristics of the Venezuelan population.

In this research, we corroborate the spontaneous recovery of most patients. Only one patient still has anosmia-type smell disorder; and in her case, other tests were carried out to establish the actual cause of such disorder. In this regard, there is similarity with what has been reported in the literature about recovery time. We did not find in our review of other investigations any relationship between the persistence of chemosensory dysfunction and the negativization of the RT-PCR test. Apparently, there is no relationship between the improvement of the disorder and the viral load detected by molecular and/or serologic tests.

The presence of asymptomatic SARS-CoV-2 positive patients and the fact that the predictive value of the Venezuelan olfactory test and the basic taste test does not exceed 70% may be hypothetically explained, not only by the time when the investigation began - which was too early with respect to the contagion peak in the country-but also by some social, demographic, and genetic characteristics of the sample studied. It could also be due to the virus mutations that affect certain populations.

In this regard, the SARS-CoV-2 spike mutation has been described as mediating the infection in human cells. Korber et al. listed 13 mutations that are deemed to be within a wider phylogenetic spectrum that could change with geography and with time, and which could give some populations certain selective advantages as to the transmission or resistance to the infection. In the case of Europe, the spike mutation D614G was dominant and began spreading in February 2020 and had such aggressive impact on the population. There is evidence of recombinant strains circulating locally and that are indicative of infections by multiple strains (22). This is being researched currently in Colombia and Venezuela. Rodríguez-Morales et al. (23) described 3 genomic sequences of coronavirus mutations that may explain the behavior of the disease in Venezuela, in specific zones like Zulia, (where there has been significant migratory activity of people to and from Colombia and Brazil) and where the severity of the infection has been higher than in other geographic zones of the country (23). This hypothesis could explain why infections are mild or asymptomatic in some areas and severe in others, or why the penetrance of chemosensory dysfunction is lower than in other reports.

We may conclude that there is significant presence of smell and taste disorders, as clinical signs, in the course of the mild coronavirus disease. Therefore, every patient who presents with symptoms, must be isolated and screened for the disease by means of appropriate diagnostic tests. Patients evolve towards the resolution of smell and taste disorders spontaneously and recover in a period of time between 3 days and 5 weeks, with an average of 8 to 10 days. Rapid diagnostic tests are not useful for diagnosing the coronavirus infection, but only for following it up. We would suggest including the olfactory test and taste test as part of the battery of tests complementary to the RT-PCR test for diagnosing SARS-CoV-2 and following up the progression of the disease.

## Data Availability

Available

## ACKNOWLEDGEMENTS

Authors acknowledge to Physicians José M. García, Francis Sánchez, Isabel Duque, David Forero, Ptte Margarett Tovar, and Ptte María Barico. Altogether contributed supporting organization, and care of patients. Special mention goes to following health Centres: Hospital Militar Universitario “Dr. Carlos Arvelo” (San Martín-Caracas), Hospital “Dr. José Ignacio Baldó” (Algodonal-Caracas), Hospital General “Dr. Jesús Yerena” (Lídice-Caracas), Hospital Oftalmológico “Dr. Francisco Risquez” (Cotiza-Caracas), Hospital de los Seguros Sociales “Dr. Miguel Pérez Carreño” (Caracas), Hospital de los Seguros Sociales “Dr. José María Vargas” (La Guaira), Hospital Militar “Nelson Sayago Mora” (La Asunción-Nueva Esparta), Hospital Militar “Dr. Vicente Salias” (Fuerte Tiuna-Caracas), Policlínica Metropolitana (Estado Miranda), Hospital Clínico Universitario de Caracas.

Other case screening centers of the Health Department of the national Armed Forces located in the Great Metropolitan Area of Caracas:

General Command of the Army and Aviation, ‘Seguros Horizonte’ headquarters, Military Prosecution Department, TVFANB headquarters, National Armed Forces Bank head office.

## AUTHORSHIP CONTRIBUTION

R. Pieruzzini, M Magris and C Ayala-Grosso, designed and performed the research. R. Pieruzzini, JJ Navas, W Rodríguez, N Parra, E Luque, A Sánchez-Gago, S González, A Hagobian, A Grullón, K Díaz, M Morales, M De Jesús, S Peña, L Rodríguez, LL Peña, A Asaro, M Magris, performed research, C Ayala-Grosso, A Hagobian, M Magris, R Pieruzzini performed research and analyzed data; R. Pieruzzini, and C Ayala-Grosso revised, analyzed, and wrote the manuscript. **R. Pieruzzini and C Ayala-Grosso** equally contributed in analyzing data and wrote the manuscript.

